# Comparison of volumetric brain analysis in subjects with rheumatoid arthritis and ulcerative colitis

**DOI:** 10.1101/2023.03.08.23286980

**Authors:** Jennifer G. Cox, Marius de Groot, Matthew J. Kempton, Steven C. R. Williams, James H. Cole

## Abstract

**Objectives:** Rheumatoid arthritis (RA) and ulcerative colitis (UC) are two autoimmune diseases where patients report high levels of fatigue, pain, and depression. The effect of systemic inflammation from these diseases is likely affecting the brain, however, it is unknown whether there are measurable neuroanatomical changes and whether these are a contributing factor to these central symptoms.

**Methods:** We included 258 RA patients with 774 age and sex matched controls and 249 UC patients with 747 age and sex matched controls in a case control study utilising the UK Biobank dataset. We used imaging derived phenotypes (IDPs) to determine whether there were differences in (1) hippocampal volume and (2) additional subcortical brain volumes between patients compared to controls and if there were common regions affected between these two diseases.

**Results:** Patients with UC had moderately smaller hippocampi compared to age and sex matched controls (difference: 134.15 mm^3^, SD ± 64.76, p = 0.035). This result was not seen in RA patients. RA patients had a significantly smaller amygdala volume than age and sex matched controls (difference: 91.27 mm^3^, SD ± 30.85, p = 0.0021, adjusted p value = 0.012). This result was not seen in UC patients. All other subcortical structures analysed were comparable between the patients and control groups.

**Conclusion:** These results indicate there are subcortical brain differences between UC, RA and controls but different regions of the limbic system are preferentially affected by UC and RA. This study may provide evidence for different neurodegenerative mechanisms in distinct autoimmune diseases.

**Key Messages:**

- Central effects such as fatigue and pain place significant burden on patients with autoimmune diseases
- Rheumatoid arthritis patients have smaller amygdala volumes compared to matched controls.
- Ulcerative colitis patients have smaller hippocampal volumes compared to matched controls.

## Introduction

Rheumatoid arthritis (RA) and ulcerative colitis (UC) are two of the most prevalent autoimmune diseases and are both projected to have increasing incidence and prevalence rates globally [1, 2]. Autoimmune diseases represent a large and heterogeneous group of disorders that afflict specific target organs [3]. However, there are common links between these various different diseases. This includes the presence of both individual and familial polyautoimmunity, which is defined as the presence of more than one autoimmune disease in a single patient or within a familial line [4].

In addition to polyautoimmunity across autoimmune diseases, there are distinct links between RA and UC specifically. A meta-analysis of published data showed that patients with UC were more than twice as likely to develop RA [5]. There is also significant overlap in the treatment of these two diseases both in the acute, flare stage and long-term disease management [6-8]. The common genetic markers and antigen patterns observed may provide some indication of shared disease pathogenesis in particular in genes related to T-cell activation and leukocyte migration [9, 10].

Traditionally, both in clinical drug development and clinical practice of autoimmune diseases, the primary focus has been in the management of disease burden in the periphery. However, the presence of central symptoms, such as pain and fatigue, present a distinct challenge in effectively treating patients with these diseases [11]. Studies have reported that 23% of UC patients suffer from depression and 33% suffer from anxiety. This has been reported to be even higher in RA with one study showing 55% of patients reporting depressive symptoms [12, 13]. In addition, RA patients with depression report increased autoimmune disease activity and lower response to treatment [7, 14].

A previous study in RA has shown smaller hippocampal volume in patients is associated with more severe functional disability and higher pain perception both on a visual analog scale and in functional pain response to pressure stimulus [15]. These central manifestations combined with increased interest in improving our understanding of the neuro-immune axis, is motivating research into whether systemic inflammation from these autoimmune pathways has a central effect. Following on from that, this raises the question whether there is a common or discrete central effect caused by different autoimmune diseases.

The aims of our study were to determine if there were structural brain differences between patients with RA compared to matched controls and patients with UC compared to matched controls. The primary analysis focused on the total hippocampal volume differences between groups with a secondary analysis looking at further subcortical regions. Additionally, we looked to see if there was overlap in the subcortical regions affected between these two diseases.

## Methods

### Study population

This is a nested case control study utilising the UK Biobank data. The UK Biobank is a large, prospective observational study of 500,000 participants providing extensive biological information [16]. The imaging substudy is planned to scan 100,000 of those participants with a standardised scanning protocol including MRI of the brain.

At the time of this investigation, brain MRI were available from 40,681 participants. For the purposes of the present study, we selected 2,028 individuals including patients with RA, UC and healthy controls. Due to the difference in age and sex distribution between RA and UC patient populations, separate controls groups were matched to each patient population in a 1:3 patient:control ratio. Using the matchit algorithm in R, an exact matching strategy was employed for sex and a nearest neighbour matching strategy was utilised for age matching and selection of the control groups. A matching ratio of 1:3 was determined to be optimal as it allowed for the highest matching ratio while utilising the matching strategy outlined above.

Data from 258 individuals with a primary or secondary diagnosis of RA identified using International Clarification of Disease (ICD)-10 codes M05 or M06 were included in the RA patient group (mean age ± SD in RA = 65.41 ± 7.06, 71% female) with 774 age and sex matched controls (mean age ± SD in RA control group = 65.41 ± 7.05, 71% female). Data from 249 individuals with a primary or secondary diagnosis of UC identified using ICD-10 code K51 were included in the UC patient group (mean age ± SD in UC = 64.06 ± 7.05, 50% female) with 747 age and sex matched controls (mean age ± SD in UC control group = 64.06 ± 7.06, 50% female).

### Data Acquisition and Processing

Full details on the UK Biobank neuroimaging data are provided here: https://biobank.ctsu.ox.ac.uk/crystal/crystal/docs/brain_mri.pdf. In Short: multi-modal MR images were acquired on a Siemens 3T scanner. The T1-weighted MRI used an MPRAGE sequence with 1-mm isotropic resolution. From the T1w data, volumetric imaging derived phenotypes (IDPs) are generated by the UK Biobank using an established image-processing pipeline [17]. The subcortical volumetric measurements specifically utilise the FMRIB Integrated Registration and Segmentation Tool (FIRST) [18].

### Statistical Analysis

All statistical analyses were carried out using R version 4.1.1.

The primary analysis for this study compared total hippocampal volume in each patient group to their respective matched control groups. Model one consisted of a linear model regression with sex, age and total intracranial volume (ICV) as covariates.

Hypertension is a known risk factor for brain atrophy and is highly associated with atrophy in the hippocampus [19]. Given this association and the increased prevalence of hypertension in both the UC and RA patient populations, as reported in the demographics table below, a second model was run analysing hippocampal volume with hypertension as an additional covariate to gender, age and ICV.

A secondary analysis utilised the remaining subcortical volume measures from the FSL FIRST pipeline and included total volume of the following structures: nucleus accumbens, amygdala, caudate, pallidum, putamen and thalamus. Multiple testing corrections for these separate measures were conducted using a Bonferroni correction across all 6 IDPs.

A tertiary analysis was completed where the individual left and right volumes were analysed in those structures where a statistically significant difference was measured in the total structure to look for potential unilateral effect.

To calculate an effect-size we used Cohen’s *d*. All p-values < 0.05 were considered statistically significant.

## Results

### Demographics

Detailed demographic information can be found in Table 1. The control groups were matched directly on age and sex. Both control groups were generally comparable to their matched patient population except for rates of hypertension and hypercholesterolemia. Hypertension specifically is nearly twice as prevalent in both patient populations as compared to their control groups. As this discrepancy was anticipated, we examine the role of hypertension in these diseases using a second model with hypertension as a covariate.

**Table 1:** Participant demographics

|  | RA<br>n=258 | RA<br>Controls<br>n=774 | p value<br>(RA vs RA<br>controls) | UC<br>n=249 | UC<br>Controls<br>n=747 | p value<br>(UC vs<br>UC<br>controls) |
| --- | --- | --- | --- | --- | --- | --- |
| Sex | 184 F /<br>74 M | 552 F /<br>222 M | 0.99 | 125F<br>/124<br>M | 375 F<br>/372 M | 0.99 |
| Age (y ± SD) | 65.41 ±<br>7.06 | 65.41 ±<br>7.05 | 0.99 | 64.06<br>± 7.05 | 64.06 ±<br>7.05 | 0.99 |
| ICV (mL) | 1155 ±<br>113 | 1160 ±<br>113 | 0.59 | 1191<br>± 112 | 1197 ±<br>116 | 0.46 |
| Hypertension % (n) | 35 (88) | 17 (128) | <0.001 | 30<br>(75) | 17 (129) | <0.001 |
| Education - % subjects<br>with a college/university<br>degree or professional<br>qualification (n) | 41 (105) | 47 (366) | 0.07 | 45<br>(113) | 48 (362) | 0.41 |
| Diabetes % (n) | 7 (17) | 4 (33) | 0.13 | 9 (22) | 5 (39) | 0.04 |
| Smoking Status |  |  |  |  |  | 0.01 |
| Current % (n) | 9 (22) | 5 (41) | 0.03 | 2 (6) | 7 (49) |  |
| Previous % (n) | 40 (105) | 36 (275) |  | 43 (106) | 35 (261) |  |
| Never % (n) | 51 (131) | 59 (458) |  | 55 (137) | 58 (437) |  |
| Ethnicity % white (n) | 95 (246) | 98 (758) | 0.03 | 95 (237) | 98 (731) | 0.03 |
| Hypercholesterolemia % (n) | 18 (47) | 9 (71) | <0.001 | 11 (28) | 9 (69) | 0.35 |

Variables listed as % (number of participants). RA stands for rheumatoid arthritis, UC ulcerative colitis, SD standard deviation, ICV intracranial volume.

### Hippocampal volume in RA and UC

A significantly lower total hippocampal volume was observed in UC patients compared to the control group (p=0.035). In contrast RA patients did not show a significant reduction in total hippocampal volume (p=0.42). A tertiary analysis was completed to investigate right and left hippocampal volumes separately to ascertain whether there was a bilateral effect. While the p value was not statistically significant for the left hippocampal volume in UC patients, the raw volume measurements combined with the calculated p value in the right (p = 0.043, Cohen’s *d* = −0.15, 95% CI −0.3, −0.01) versus left (p = 0.096, Cohen’s *d* = −0.13, 95% CI −0.27, 0.02) hippocampal volume differences are however similar enough not to suggest lateralization of the difference observed. The results for total hippocampal volume in UC can be found in Fig. 1. Full results of the primary and tertiary analysis in hippocampal volumes for both UC and RA can be found in Table 2.

**Table 2:** Hippocampal volume in RA and UC

|  | Region of Interest | p-value | Mean Volume Patient Group (mL) | Mean Volume Control Group (mL) | Cohen's *d* | 95% CI |
| --- | --- | --- | --- | --- | --- | --- |
| RA | Left Hippocampus | 0.68 | 3.7 +/- 0.4 | 3.7 +/- 0.4 | -0.04 | -0.18, 0.1 |
|  | Right Hippocampus | 0.31 | 3.8 +/- 0.5 | 3.8 +/- 0.5 | -0.08 | -0.22, 0.06 |
|  | Total Hippocampus | 0.42 | 7.5 +/- 0.9 | 7.5 +/- 0.8 | -0.07 | -0.21, 0.07 |
| UC | Left Hippocampus | 0.096 | 3.7 +/- 0.5 | 3.8 +/- 0.5 | -0.13 | -0.27, 0.02 |
|  | **Right Hippocampus** | **0.043** | **3.8 +/- 0.5** | **3.9 +/- 0.5** | **-0.15** | **-0.3, -0.01** |
|  | **Total Hippocampus** | **0.035** | **7.6 +/- 0.9** | **7.7 +/- 0.8** | **-0.16** | **-0.3, -0.01** |

Regions with significant findings shown in bold.

**Fig. 1.**
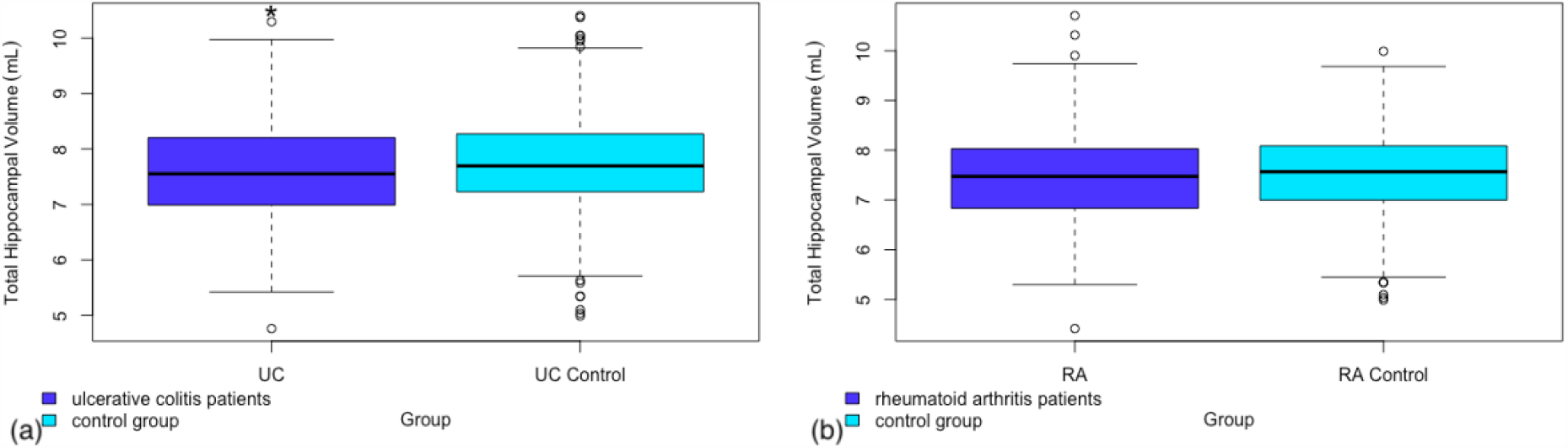
Box and whisker plot representing the total hippocampal volume in (a) patients with ulcerative colitis as compared to a matched control group (b) patients with rheumatoid arthritis as compared to a matched control group

As mentioned in the methods section, a second model was run including hypertension as a covariate. This was performed to account for any potential signal being attributable to the most prevalent cerebrovascular risk factor in these patient populations. This did account for some of the signal with a p-value of 0.08 in model 2 for total hippocampal volume in UC versus controls compared to 0.035 in model 1 without hypertension as a covariate. Full results can be found in Table 5 in the supplementary material.

### Additional subcortical regions analysis

A secondary analysis was performed in both RA and UC patients looking at total volume of additional subcortical regions provided by the FSL FIRST pipeline. Of the additional regions analysed there was a significant difference in amygdala volume between RA patients and controls (Figure 2). No other sub-cortical regions showed significant volume differences between RA patients and controls (see Table 3).

**Table 3:** Additional Subcortical Regions RA

| Region of Interest | p-value | Adjusted p-value | Mean Volume RA (mL) | Mean Volume Controls (mL) | Cohen's *d* | 95% CI |
| --- | --- | --- | --- | --- | --- | --- |
| Accumbens | 0.22 | 0.99 | 0.8 +/- 0.2 | 0.9 +/- 0.2 | -0.09 | 0.23, 0.05 |
| **Amygdala** | **0.0021** | **0.012** | **2.4 +/- 0.4** | **2.5 +/- 0.4** | **-0.21** | **-0.35, -0.07** |
| Caudate | 0.43 | 0.99 | 6.8 +/- 0.8 | 6.8 +/- 0.8 | 0.02 | -0.12, 0.16 |
| Pallidum | 0.52 | 0.99 | 3.4 +/- 0.5 | 3.5 +/- 0.5 | -0.06 | -0.2, 0.08 |
| Putamen | 0.44 | 0.99 | 9.3 +/- 1.2 | 9.3 +/- 1.1 | 0.02 | -0.12, 0.16 |
| Thalamus | 0.88 | 0.99 | 14.8 +/- 1.5 | 14.9 +/- 1.5 | -0.04 | -0.18, 0.11 |

Regions with significant findings shown in bold; adjusted p value includes a Bonferroni multiple testing correction

**Figure 2:**
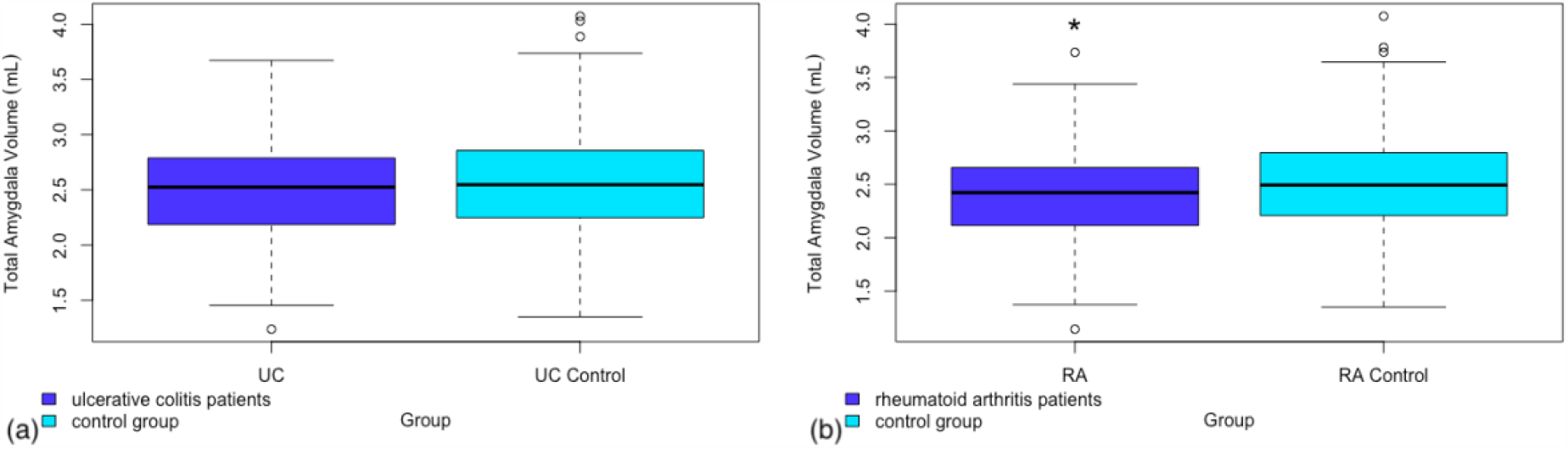
Box and whisker plot representing the total amygdala volume in (a) patients with ulcerative colitis as compared to a matched control group (b) patients with rheumatoid arthritis as compared to a matched control group.

This same result was not found in the UC population. There was no significant difference seen between UC patients and controls in any of the additional subcortical regions analysed. Full results from this analysis can be found in Table 4.

**Table 4:** Additional subcortical regions UC

| Region of Interest | p-value | Adjusted p-value | Mean Volume UC (mL) | Mean Volume Controls (mL) | Cohen's *d* | 95% CI |
| --- | --- | --- | --- | --- | --- | --- |
| Accumbens | 0.12 | 0.74 | 0.9 +/- 0.2 | 0.9 +/- 0.2 | -0.11 | -0.26, 0.03 |
| Amygdala | 0.057 | 0.34 | 2.5 +/- 0.4 | 2.6 +/- 0.5 | -0.15 | -0.29, 0 |
| Caudate | 0.78 | 0.99 | 6.9 +/- 0.8 | 7.0 +/- 0.8 | -0.05 | -0.19, 0.09 |
| Pallidum | 0.95 | 0.99 | 3.6 +/- 0.5 | 3.6 +/- 0.5 | -0.02 | -0.17, 0.12 |
| Putamen | 0.51 | 0.99 | 9.6 +/- 1.1 | 9.6 +/- 1.1 | 0.01 | -0.14, 0.15 |
| Thalamus | 0.90 | 0.99 | 15.2 +/- 1.4 | 15.3 +/- 1.5 | -0.03 | -0.18, 0.11 |

## Discussion

This is the largest dataset, to date, investigating subcortical brain volume measures in both RA and UC. In patients with UC compared with controls, we found a moderately smaller total hippocampal volume. Additional analyses were conducted looking at the right and left hippocampus separately and the resulting p-values did not indicate a strong unilateral affect. No similar, significant reduction in hippocampal volume was observed in the RA population.

In the secondary analysis, we found a significantly lower amygdala volume in patients with RA compared to controls. This same result was not seen in the UC population. These findings may suggest there are different areas of the brain effected in these two diseases. At the same time, the close relation between the amygdala and hippocampus may also suggest a common pathway affecting the brain in these diseases. This is particularly interesting as both the amygdala and the hippocampus are associated with chronic pain and depression [20, 21]. However, the fact that both regions are not affected in the same way in these two diseases suggests potential differences in the specific disease processes that may preferentially affect one region over another and may result in varying degrees of these central symptoms. The significant difference in amygdala volumes in RA may be linked to the increased rates of anxiety in RA versus UC patients. Previous rs-fMRI research has suggested that the cognitive impairments and reported “brain fog” in UC may be predominantly linked to the limbic system which would be consistent with these findings [22].

Increased rates of cardiovascular risk factors are a known complication of autoimmune diseases. The prevalence of hypertension seen in our RA population is consistent with previously published literature [23]. In the case of UC, it is more complicated with some of the literature reporting higher rates of hypertension and some reporting similar rates of hypertension to the general population, but higher rates of other cardiovascular risk factors [24]. In both diseases there is extensive literature pointing to a host of higher cardiovascular risk factors with unknown aetiology [25]. It has been proposed that this could be a result of medication usage, lack of physical activity due to disease burden and/or a function of the disease process directly [26, 27]. Irrespective of the origin of these risk factors there is clear evidence of localised, lower subcortical volumes in both diseases.

One limitation to this study is the lack of information on disease duration and severity. Both UC and RA are relapsing-remitting diseases and can vary widely in their individual presentation. In a recent paper Zhang et al. sub-categorised UC patients into active stage versus remission and reported fewer regions of decreased neuroanatomical volume in those currently in remission [28]. There is also previously published data that suggests that the age of onset can significantly affect disease severity [29]. How well controlled an individual’s disease is and on what medications may also have an important impact on these central effects which we are unable to quantify with this data set.

An additional limitation of this work is the lack of longitudinal data. There is an ongoing project to acquire follow up scans in a subset of up to 10,000 UK Biobank Imaging cohort participants. This may make it possible to analyse atrophy over time in these patient populations, but it is unclear how many of the patients with an RA or UC diagnosis will be included in that follow up study.

One ongoing point of debate with regards to the UK Biobank data set is the lack of heterogeneity and the question of whether it is truly representative of the wider population. Participants tend to be healthier than the general population reporting lower rates of cancer and overall all-cause mortality in addition to being more health conscious. This is a well-established effect seen in volunteer-based cohort studies [30]. While this is important to acknowledge, the breadth of lifestyle, genetic and demographic information allows us to appropriately contextualise our patient and control populations. As a resource this allowed us access to neuroimaging data in these two diseases that is substantially larger than anything published to date.

These findings play a potentially important role in further understanding brain volume differences in preferential areas of the brain in RA and UC. With increased focus on understanding and mitigating risk factors for neurodegenerative diseases this provides the potential foundation for future work in exploring the link between these autoimmune diseases and the development of future neurodegenerative diseases in these populations. There is a known link between UC and Parkinson’s Disease (PD) and between RA and Alzheimer’s Disease (AD). There is also documented research suggesting accelerated atrophy in PD in the hippocampus, as seen in our results for UC, and in the amygdala in AD, as seen here in our RA results [31, 32]. This highlights the importance of continued monitoring and treatment of central symptoms for brain health long term in people with chronic autoimmune conditions.

## Data Availability

All data produced in the present work are contained in the manuscript

## Acknowledgements

From the Department of Neuroimaging and the Department of Psychosis Studies, Institute of Psychiatry, Psychology, and Neuroscience, King’s College London

## Data Availability

UK Biobank data are available through a procedure described at https://www.ukbiobank.ac.uk/enable-your-research. All data was accessed under UK Biobank application number 40933.

