## Supplemental Table for "Comparison of volumetric brain analysis in subjects with rheumatoid arthritis and ulcerative colitis"

Supplementary Materials for "Comparison of volumetric brain analysis in subjects with rheumatoid arthritis and ulcerative colitis"

Table 5: Model 2 for hippocampal volume in RA and UC includes gender, age, ICV and hypertension as covariates

| | Region of Interest | p-value | Mean Volume Patient Population (mL) | Mean Volume Control Population (mL) | Cohen's d | 95% CI |
|---|---|---|---|---|---|---|
| RA | Left Hippocampus | 0.84 | 3.7+/- 0.5 | 3.7 +/- 0.4 | -0.04 | -0.18, 0.1 |
| | Right Hippocampus | 0.36 | 3.8 +/- 0.5 | 3.8 +/- 0.5 | -0.08 | -0.22, 0.06 |
| | Total Hippocampus | 0.52 | 7.5 +/- 0.9 | 7.5 +/- 0.8 | -0.07 | -0.21, 0.07 |
| UC | Left Hippocampus | 0.20 | 3.7 +/- 0.5 | 3.8 +/- 0.5 | -0.13 | -0.27, 0.02 |
| | Right Hippocampus | 0.08 | 3.8 +/- 0.5 | 3.9 +/- 0.5 | -0.15 | -0.3, -0.01 |
| | Total Hippocampus | 0.08 | 7.6 +/- 0.9 | 7.7 +/- 0.8 | -0.16 | -0.3, -0.01 |
